# Patient and family reported clinical picture of *IRF2BPL*-related disorders

**DOI:** 10.64898/2026.03.03.26347377

**Authors:** Zoe Goldstone-Joubert, Danielle M. Pascual, Laurie Anne Bailey, Loren D.M. Peña, Paul C. Marcogliese

## Abstract

IRF2BPL-related disorder is a neurodevelopmental disorder caused by heterozygous variants in the *IRF2BPL* (Interferon Regulatory Factor 2 Binding Protein-Like) gene. The few reports available in the literature suggest that common symptoms include developmental delay, intellectual disability and developmental regression. There are no reports of genotype-phenotype correlations. We developed a retrospective and prospective patient-reported survey to assess diagnostic information, presenting symptoms and longitudinal follow-up of neurological symptoms for up to two years. Clinical information was available for all 32 participants and was highly variable in regards to age at symptom onset, severity of neurologic manifestations, and progressivity. For 27 of the 32 participants, diagnostic genetic test results were available. Genetic mutation analysis revealed 22 individuals with truncating variants and five participants with unique missense variants in *IRF2BPL*. The study data support the hypothesis that *IRF2BPL* missense variants are associated with a less severe disease presentation and progression than participants with truncating variants. The purpose of this study is to further define IRF2BPL-related disorder and provide more clinical and molecular insight into this ultra-rare disease.

**Highlights:**

- Patient-reported clinical history at diagnosis and up to two years of follow up
- The clinical spectrum is increasingly heterogeneous
- We report 32 patients, 27 with noted *IRF2BPL* variants, 14 being novel to literature.
- Data supports the notion that *IRF2BPL* missense variants may be associated with less severe disease than truncations (nonsense/frameshifts).

## Introduction

The establishment of a molecular diagnosis for rare genetic disease has been made possible by the identification of novel gene variants using a combination of next-generation sequencing, control databases, match-making networks, and variant functional testing. This has been particularly significant for rare Mendelian disease, many of which are neurodevelopmental disorders (NDDs). In 2018, the Undiagnosed Disease Network^1^ and their collaborators identified the first cohort of patients with heterozygous, *de novo* alterations in the single-exon gene *IRF2BPL* (Interferon Regulatory Factor 2 Binding Protein-Like) as the cause of a severe NDD^2^. The disorder was named NEDAMSS (neurodevelopmental disorder with regression, abnormal movements, loss of speech, and seizures) – MIM#618088^3^. Since the initial cohort, a stream of additional cases have been reported with over 80 disease-associated variants now published^4–23^. However, as with other NDDs, the increase in identified genetic variants is associated with increasing variability of symptoms, age of onset, and symptom severity without any obvious genotype-phenotype correlations. Reports suggest that individuals with autosomal dominant changes in *IRF2BPL* display a range of symptoms that usually includes some typical development and acquisition of skills, including speech, that may be followed by regression that can be progressive. Although regression occurs on average, around five years of age, it has been observed as early as eight months and up to young adults over 20 years old^7,14^.

The majority of reported patients have some degree of global developmental delay with and without seizures. The seizure type is not consistent between patients and includes generalized tonic-clonic, myoclonic, absence, focal tonic-clonic, complex partial, infantile spasms, and/or atonic seizures^24^. Other reported neurologic symptoms include hypotonia, dystonia, choreoathetosis, ataxia, dysmetria, dysarthria, and dysdiadochokinesia. Additionally, a significant portion of patients are reported to display eye movement abnormalities, such as gaze-evoked nystagmus and limited upward gaze^9^. Additional reported eye findings include dysconjugate gaze, ophthalmoplegia, gaze palsy, and slow saccades. Rare ocular features include keratoconus^13,14^, cataracts^13^, retinal pigmentary anomalies^10^, and macular degeneration in a single individual^2^. Interestingly, cerebellar and cerebral white matter atrophy have been observed in patients undergoing MRI^9^ and a recent case report included the first published post-mortem analysis showing cortical, cerebellar and nigral degeneration^26^. Notably, rare variants in *IRF2BPL* have also been observed in larger populations of patients with early-onset parkinsonism^27^. Some patients with missense variants have been observed to display milder neurological symptoms including autism spectrum disorder (ASD) and rare *de novo* variants in *IRF2BPL* have been implicated in larger populations of people with ASD^25^. Although reported, non-neurological symptoms such as facial dysmorphia and hypotonia, joint contractures, and cardiomyopathy are less common^17^.

As more clinicians order exome and whole genome sequencing to attempt to identify a molecular explanation for patient symptoms it is expected that many more individuals with pathogenic *IRF2BPL* changes will be identified. Although evaluating more patients is helpful in determining the full breadth of clinical manifestations of IRF2BPL-related conditions, it is still an ultra-rare disease. As rare diseases impose a significant reduction in the quality of life of both the patient and their families, it is important to note that the patients and families who take care of the affected are central to learning more about their everyday symptoms and experiences. Patient-reported outcome measures (PROMs) capture patients’ perspectives on their health, which can help us gain more insight into their quality of life and representative symptoms^28^. No PROM has been created specifically for the IRF2BPL community. To address this issue, we designed a PROM REDCap survey to collect IRF2BPL patient/parent-reported clinical manifestations. Data was collected at the time of consent, and in a subset of patients, neurologic data was collected every 6 months for up to 2 years. The purpose of this survey and the patient registry is to provide valuable insights into the natural history of NEDAMSS while also encouraging families to actively participate in research. This effort aims to enhance our understanding of the variability of the clinical manifestations and assess the disease’s impact on patients, caregivers, and quality of life. Genetic diagnostic information was also collected. Attempts will be made to align phenotypic disease presentations with functional variant categories, ie missense mutations versus truncating (nonsense and frameshift) mutations.

## Material and methods

### Instrument

A retrospective parent-reported online questionnaire was developed via REDcap, an online survey tool, to assess the natural history of patients with IRF2BPL-related disorder. The questionnaire was broken into three surveys to obtain 1) demographic and diagnostic information, 2) initial neurological symptoms, and 3) follow-up neurological symptoms. Follow-up neurological symptom surveys were distributed every six months upon recruitment into the study, with follow-up data obtained up to 24 months post-study recruitment. The study is currently closed to recruitment and no additional data is being collected.

### Participants and Recruitment

Patients diagnosed with IRF2BPL-related disorder were invited to participate in a retrospective parent-reported online questionnaire. This study was reviewed and approved by the Cincinnati Children’s Hospital Institutional Review Board (protocol 2023-0410). Participants were recruited via posting on the ClinicalTrials.gov website, with advocacy groups for NEDAMSS, the Undiagnosed Diseases Network’s page for IRF2BPL, and contact from interested families and providers. All patients with a genetic diagnosis of an IRF2BPL-related disorder were eligible for the study. Parents or guardians were asked to complete the questionnaire for patients less than 18 years of age or if they were severely impaired. For patients with milder phenotypes, a study investigator assessed the participant’s ability to provide consent. Participants were explained the purpose of the registry and were subsequently consented. After consent was obtained, participants’ parents or legal guardians were asked to submit documentation of diagnosis, which was reviewed by a study investigator. If the participant received care at the CCHMC, records were obtained through the participant’s medical record. 34 participants consented to participate in the study and 32 participants completed at least the initial survey.

### Data Collection

Participants submitted answers to a total of five surveys through REDcap. The initial two surveys included demographic or diagnostic information and neurological symptoms. The subsequent surveys consisted of follow-up neurological information that was filled out at 6, 12, 18 and 24 months. It should be noted that although patient-reported data is increasingly being used in research and may give new perspectives in terms of quality of life – the accuracy of responses obtained from participants have not neccessarly involved collaboration of clinicians. Moreover, since data is anonymized, data herein may overlap with previously published case reports.

### Data Analysis

Data was extracted from REDcap^29^, analysis and graphical representation was completed in GraphPad Prism version 10.

## Results

### Patient Demographics

32 participants completed the initial demographic survey which consisted of general demographic questions and questions regarding diagnostic information. Of the 32 participants, 84% (27/32) identified as European ancestry, 6% (2/32) identified as African or African American and 9% (3/32) identified as East Asian (**Fig. 1A**). Age of diagnosis ranged from under one years of age to over 18 years of age, with the most common ages of diagnosis being between one to five years of age (34.4%) followed by 11 to 17 years of age (25%) (**Fig. 1B**). Genetic variants in the *IRF2BPL* gene were identified using four different methods including whole-genome sequencing (WGS), whole-exome sequencing (WES), panel testing and single gene sequencing, with WES being to most common method of diagnosis (74%) (**Fig. 1C**, Supplemental Table 1). The majority of genetic variants identified were truncating variants (68.8%, 22/32) including nonsense variants (34.4%) and frameshift variants (34.4%). Missense variants were found in a minority of participants (15.6%, 5/32). In total, 15.6% (5/32) of participants did not know the type of genetic change (**Fig. 1D**). There were a number of symptoms that prompted genetic testing including fine motor delay, speech delay, gross motor delay, loss of developmental milestones, seizures, hypotonia, abnormal movements, and autism (**Fig. 1E**).

**Fig. 1.**
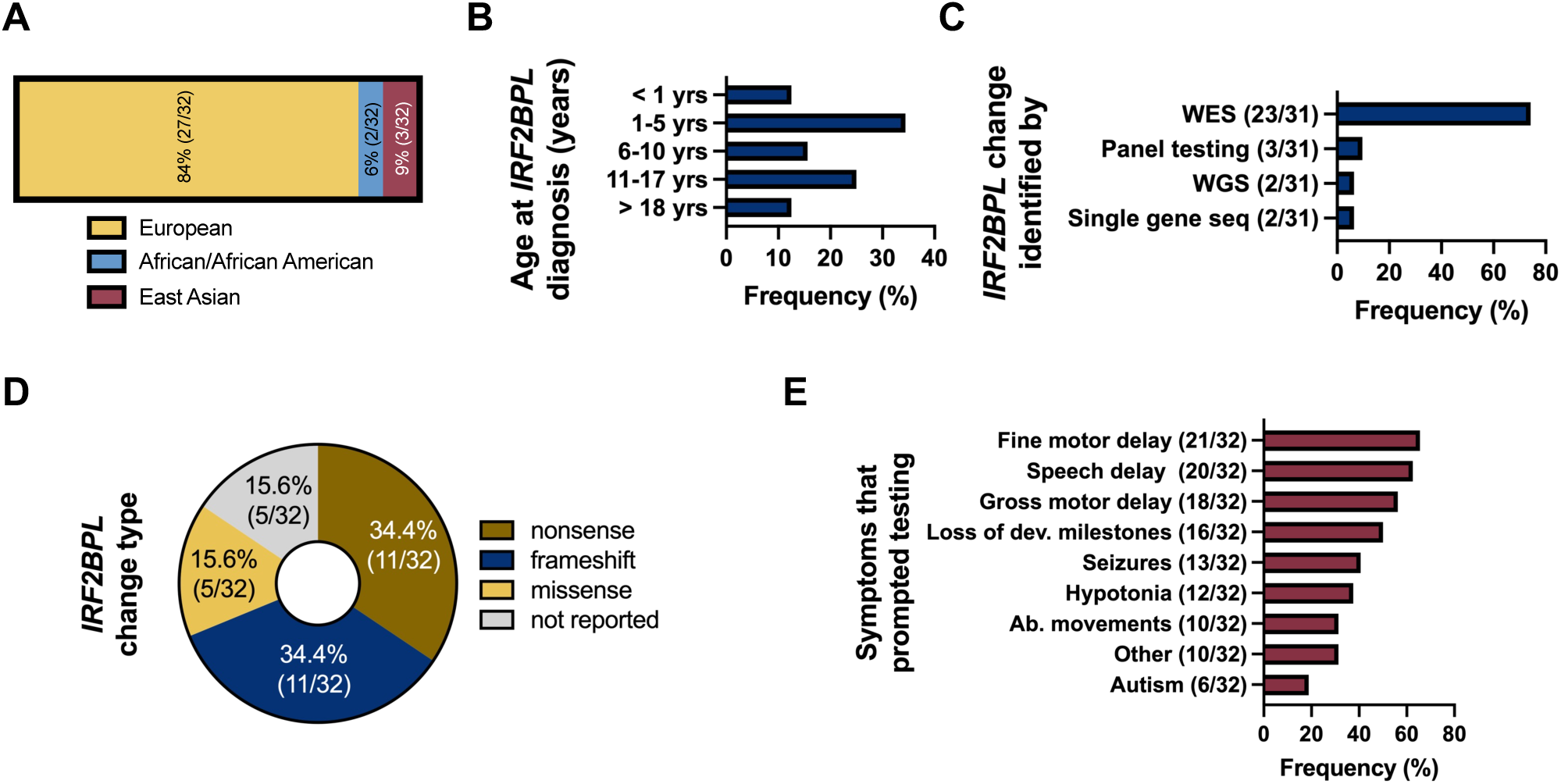
Study demographics and referral information. This includes the self-identified population of patients (A) and age of IRF2BPL diagnosis (B). The genetic alteration in IRF2BPL was identified by WES (whole-exome sequencing), Panel testing including IRF2BPL, WGS (whole-genome sequencing), or single gene seq (sequencing) for IRF2BPL (C). Variant class of IRF2BPL change was asked of respondents (D). List of symptoms present in patients that promoted genetic testing (E).

### Symptoms at Study Onset

Participants completed an initial survey to assess overall symptoms including gross motor milestones, language milestones, self-help skills, and developmental regression. The survey also asked questions regarding types of developmental services frequented by patients. For overall symptoms (**Fig. 2A**), the most prominent feature was developmental delay (DD), reported in 81.3% (26/32) of participants. Intellectual disability (ID) and developmental regression (DR) were reported in roughly 50% (15/32) of participants. Other symptoms included seizures (40.6%, 13/32), abnormal movement of the extremities (37.5%, 12/32), symptoms of autism (31.3%, 10/32), swallowing issues (31.3%, 10/32), abnormal brain MRI (21.9%, 7/32), feeding intolerances (18.8%, 6/32), joint contractures (9.4%, 3/32), and respiratory support (6.3%, 2/32). Other than joint contractures and respiratory support, all symptoms listed above were seen in both patients with truncations and missense variants.

**Fig. 2.**
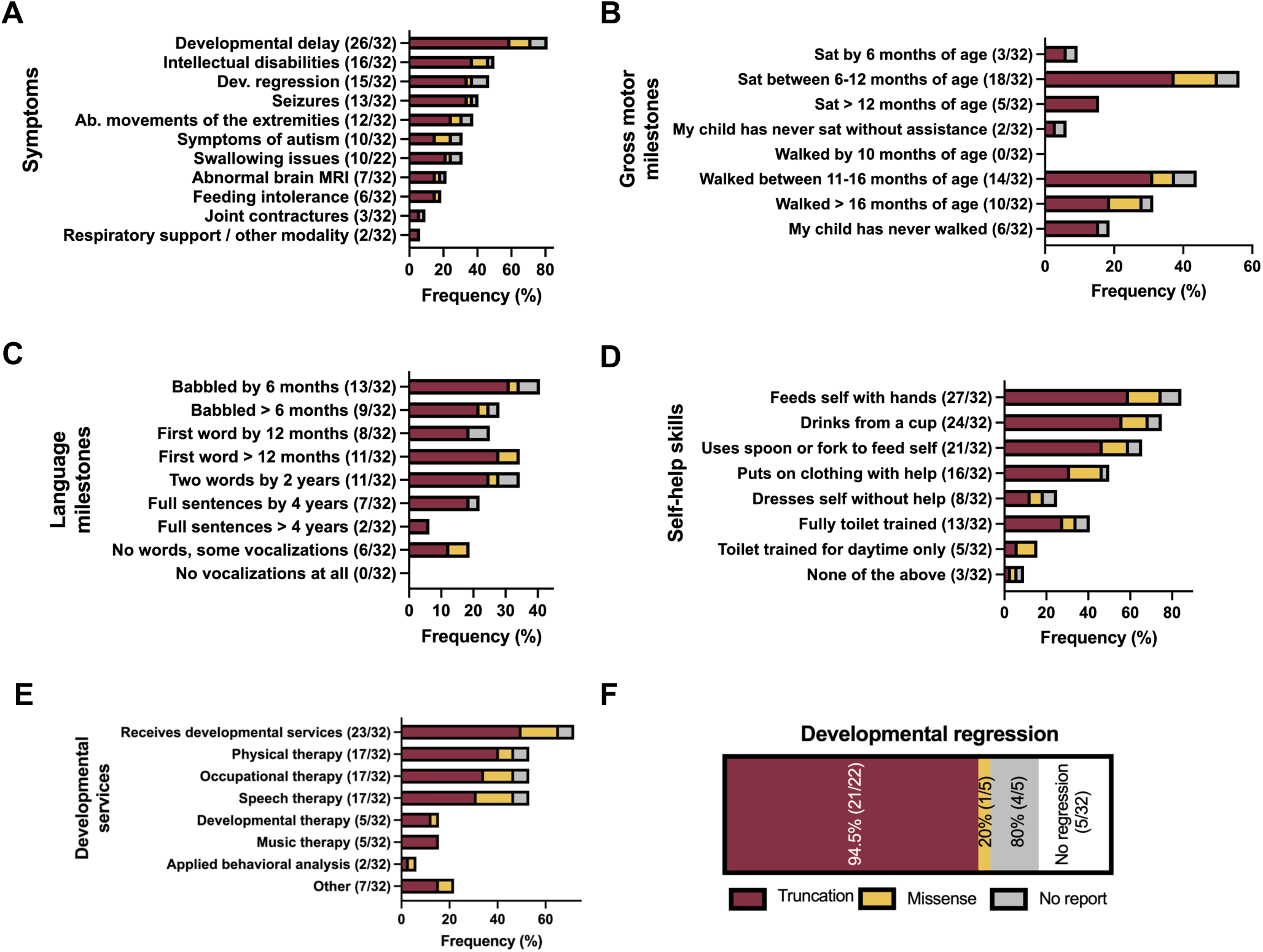
Symptomology of IRF2BPL-related patients at study onset. Overall symptoms (A), gross motor (B) and language (C) milestones, self-help skills (D), presence of regression (E), and use of developmental services (F). Burgundy = truncation (nonsense or frameshift), yellow = missense, grey = no genetics reported.

Gross motor milestones were assessed based on age of sitting and walking (**Fig. 2B**). Most patients (56.3%, 18/32) first sat between six and 12 months of age. Most patients (43.8%, 14/32) walked between 11 and 16 months of age. Some patients never sat without assistance (6.25%, 2/32) and some have yet to walk independently (18.8%, 6/32). Patients with missense variants were not part of either group that never sat or walked independently.

Language milestones were assessed based on age when the patient first babbled, when first and second words were spoken and when the patient developed full sentences (**Fig. 2C**). For babbling, 40.6% (13/32) of patients babbled by 6 months whereas 28.1% (9/32) babbled after 6 months. In 25% (8/32) of patients, their first word was spoken before 12 months and 34.4% (11/32) was after 12 months. For second words, 34.4% (11/32) were able to speak two words by 2 years of age. For full sentences, 21.9% (7/32) of patients were able to speak full sentences by 4 years 6.3% (2/32) were able to speak full sentences after 4 years. In 18.8% (6/32) of patients, only vocalizations were developed.

Self-help skills were divided into different groups and assessed based on patients’ abilities (**Fig. 2D**). For eating abilities, most of the patients were able to feed themselves with their hands (84.4%, 27/32), drink from a cup (75%, 24/32) and use a spoon or fork to feed themselves (65.6%, 21/32). For dressing abilities, 50% (16/32) of patients were able to put clothing on with help whereas only 25% (8/32) of patients were able to dress themselves independently. For toilet training abilities, a minority of patients (15.6%, 5/32) were toilet trained during the daytime only and roughly half of patients (49.6%, 13/32) were fully toilet trained. In 9.4% (3/32) of patients, none of the skills mentioned above were acquired. The level of self-help skills was spread out evenly among patients with truncating or missense variants.

It was assessed if patients received developmental services and further broken down into the types of services received (**Fig. 2E**). The majority of patients (71.9%, 23/32) received some type of developmental service. Of the different types of services, the most common services included physical therapy, occupational therapy and speech therapy for with 53.1% (17/32) of patients utilized. Fewer patients received developmental therapy (15.6%, 5/32), music therapy (15.6%, 5/32), and applied behavioural analysis (6.3%, 2/32).

DR was examined more closely (**Fig. 2F**). Of the patients with truncating variants, 94.5% (21/22) displayed DR in either gross motor, fine motor, language, or self-help skills. For patients with missense variants, only 20% (1/5) displayed DR. In 15.6% (5/32) of patients, no DR was reported. For 16 of the patients with DR, 37.5% (6/16) displayed difficulties sitting or walking as the initial symptom of regression, 25% (4/16) started to stumble or fall more than usual, 18.8% (3/16) had difficulty with articulation, 6.3% had loss of spoken language and 12.5% (2/16) had other initial symptoms (**Fig. 3A**).

**Fig. 3.**
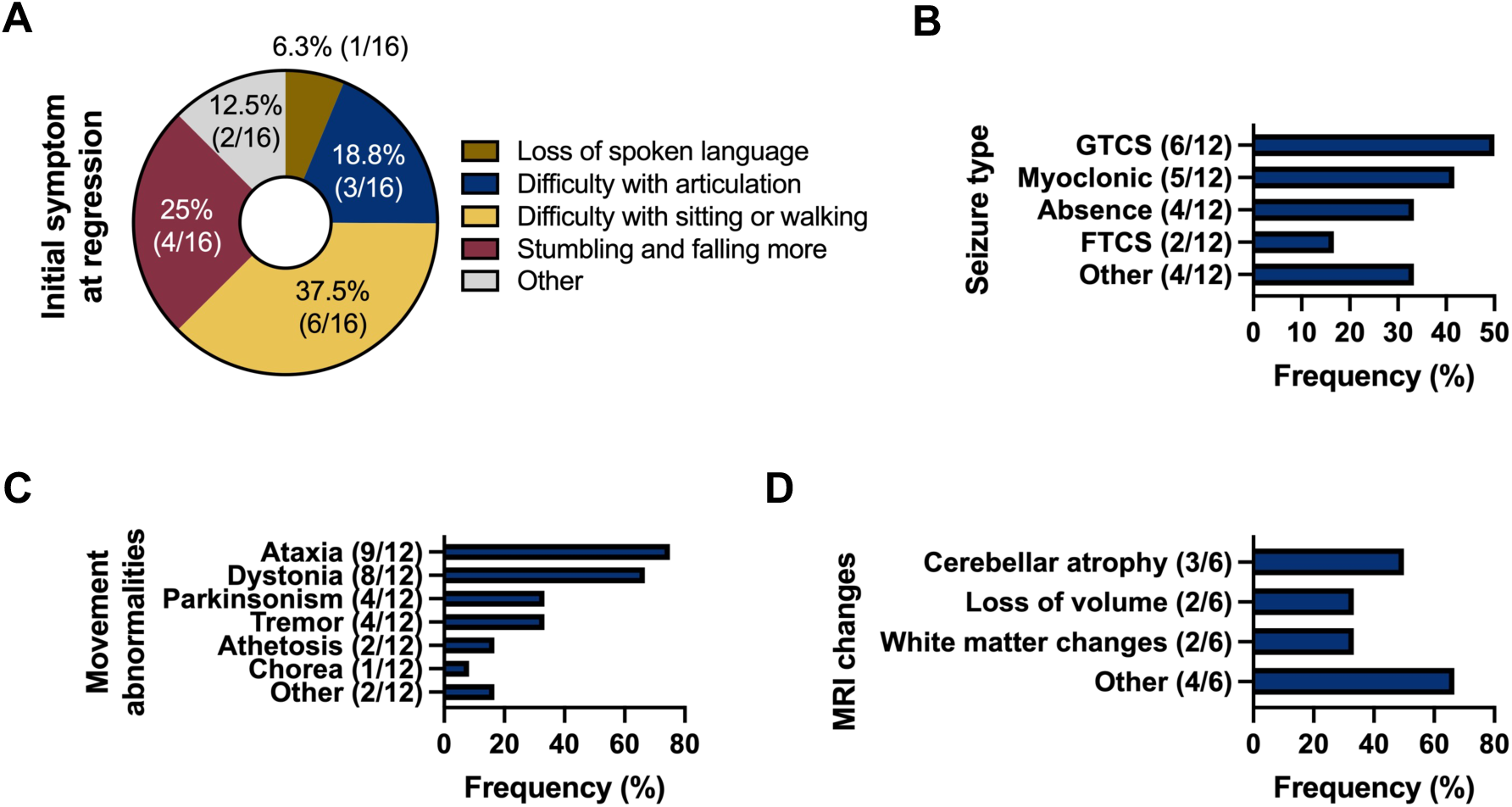
Neurological details of IRF2BPL-related patients. Respondents were asked what were the initial symptoms at developmental regression (A), seizure type, movement abnormality details, and any imaging changes on MRI. GTCS = generalized tonic-clonic seizures, FTCS = focal tonic-clonic seizures.

### Initial Neurological Symptoms

Initial neurological symptoms were examined and broken up into different categories including seizure type, movement abnormalities and MRI changes. Seizures of various types were seen in 37.5% (12/32) of patients including generalized clonic seizures (GTCS) (50%. 6/12), myoclonic seizures (41.7%, 5/12), absence seizures (33.3%, 4/12), focal tonic-clonic seizures (FTCS) (16.7%, 2/12) or other seizure types (33.3%, 4/12) (**Fig. 3B**). Movement abnormalities were also reported in 37.5% (12/32) patients and included ataxia (75%, 9/12), dystonia (66.7%, 8/12), Parkinsonism (33.3%, 4/12), tremors (33.3%, 4/12), athetosis (16.7%, 2/12), chorea (8.3%, 1/12) or another type of movement disorder (16.7%, 2/12) (**Fig. 3C**). MRI changes were reported in fewer patients (18.8%, 6/32). MRI changes included cerebellar atrophy (50%, 3/6), loss of brain volume (33.3%, 2/6), white matter changes (33.3%, 2/6) or other changes (66.7%, 4/6) (**Fig. 3D**).

### Neurological Symptoms During the Two-Year Follow-Up Period

A subset of patients (16/32) participated in follow-up surveys regarding seizure, motor, imaging, speech and ocular details every six months throughout a two-year period. Over the course of six months, 43.8% (7/16) of patients had an electroencephalogram (EEG) in the past year compared to 14.3% (1/7) at 24 months (**Fig. 4A**). Of the patients who had an EEG, abnormal results were seen at all follow-up periods except 24 months. It was found that a minority (6.3 to 14.3%) of patients had new seizure onset at some point during the 24-month follow-up timeframe. Of the patients who developed seizures during the follow-up period, seizure types included GTCS, absence, myoclonic or unknown (**Fig. 4B**). Over the course of 24 months, the onset of new movement abnormalities was seen more frequently than new seizure onset and ranged from 11.1% to 42.1% of patients (**Fig. 4C**). Commonly seen motor abnormalities included ataxia, dystonia and tremors (**Fig. 4D**). Few patients (7.7% to 18.8%) received new brain imaging in the 24-month follow-up period (**Fig. 4E**). Abnormal MRI findings included loss of brain volume, brain atrophy, cerebellar atrophy, or other changes (**Fig. 4F**). In comparison to new seizure and movement abnormality onset, more patients (28.8% to 56.3%) developed speech challenges or existing speech challenges worsened over the course of 24 months (**Fig. 5A**). Patients developed a variety of speech challenges including aphasia, oral apraxia, dysphasia, or other challenges (**Fig. 5B**). Over the course of 18 months, some patients (12.5% to 38.5%) developed new ocular changes (**Fig. 5C**) which consisted of strabismus, disconjugate gaze, nystagmus, or other changes (**Fig. 5D**). At 24 months of follow-up, none of the seven patients who participated reported new ocular changes.

**Fig. 4.**
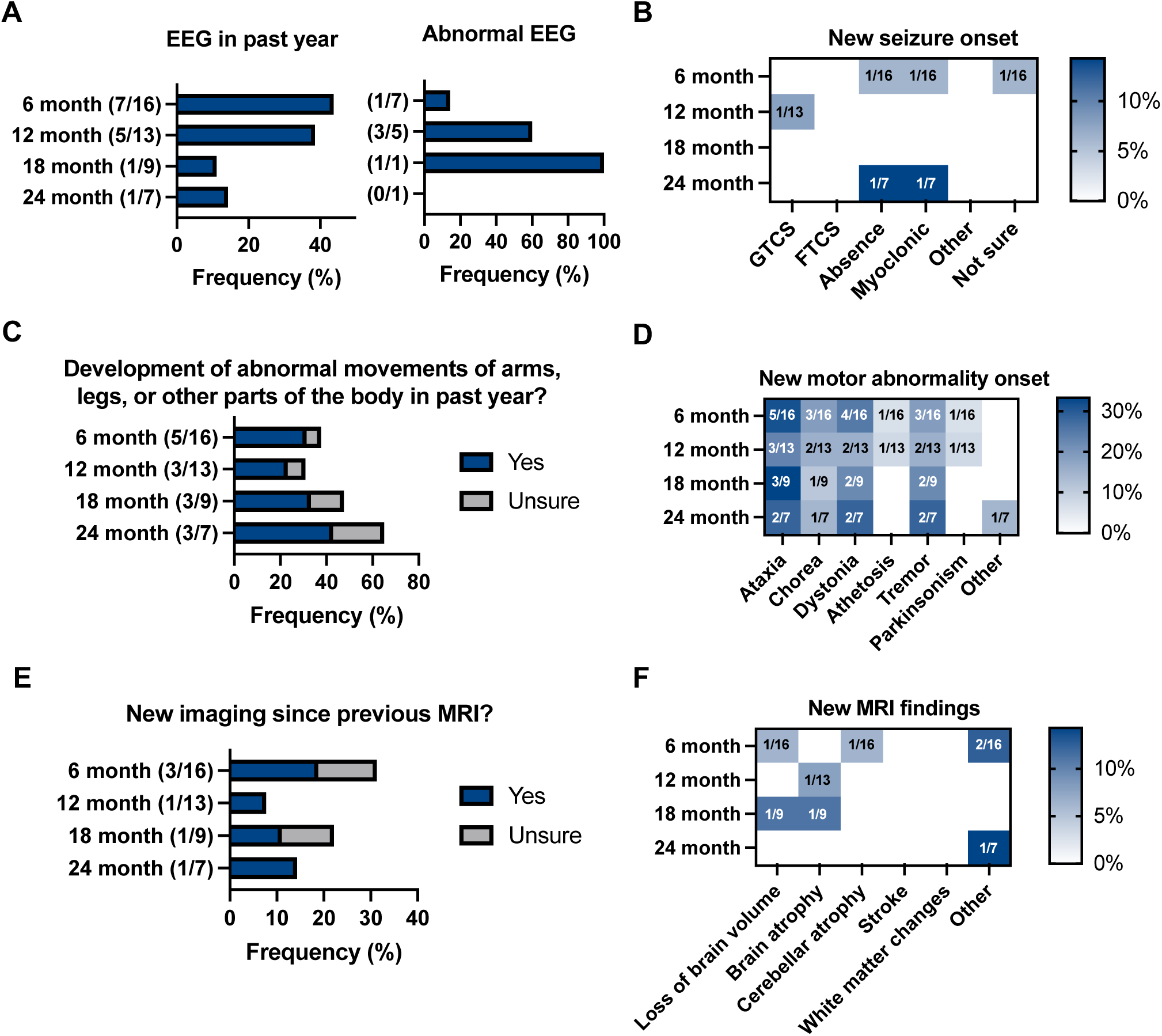
Seizure, motor, and imaging details for two-year neurological follow-up. Respondent information for 6, 12, 18, and 24 month neurological follow up including EEG (electroencephalogram) (A), new seizure type details (B), development of abnormal motor phenotypes (C) across follow-up reports (D), and new MRI studies (E) with findings (F). GTCS = generalized tonic-clonic seizures, FTCS = focal tonic-clonic seizures.

**Fig. 5.**
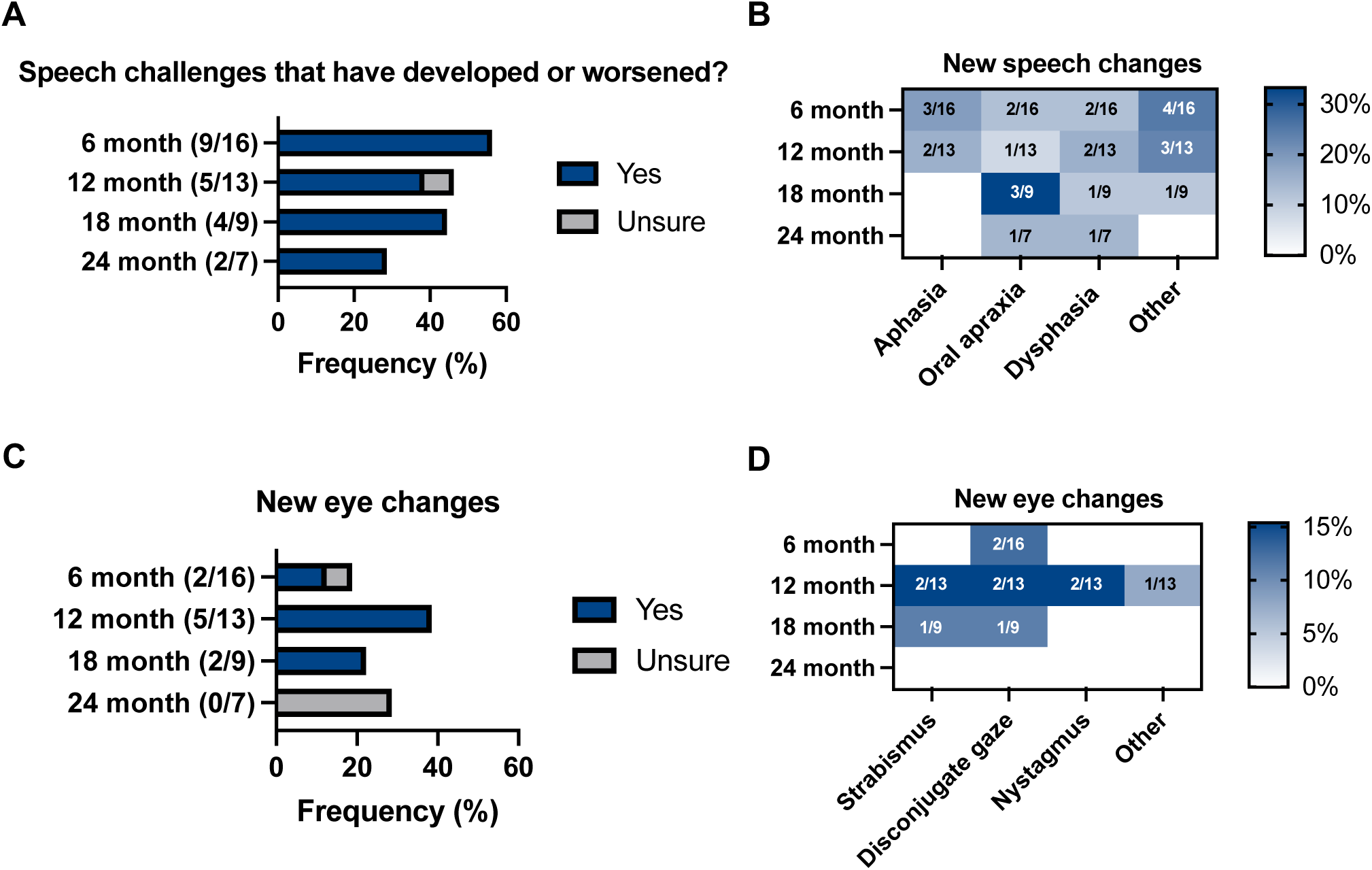
Speech and ocular details for two-year neurological follow-up. Respondent information for 6, 12, 18, and 24 month neurological follow-up speech challenges (A) and whether patients develop aphasia (complete loss of speech), oral apraxia (difficulty with specific sounds), or dysphagia (difficulty understanding patient’s speech) (B), development of eye phenotypes (C) across follow up reports (D).

### Variant Segregation

Truncating variants can be seen throughout the *IRF2BPL* gene. However, they tend to cluster in the poly alanine (pA) and poly glutamine (pQ) tracts. In addition, truncating variants demonstrate a wide clinical heterogeneity, independent from the region they are found in the gene. With few missense variants, it is difficult to determine if these variants cluster in certain regions of the gene. However, there seems to be a cluster in the middle of the gene from the 350 to 500 base pair region. Like truncating variants, missense variants also display a large clinical heterogeneity (**Fig. 6**).

**Fig. 6.**
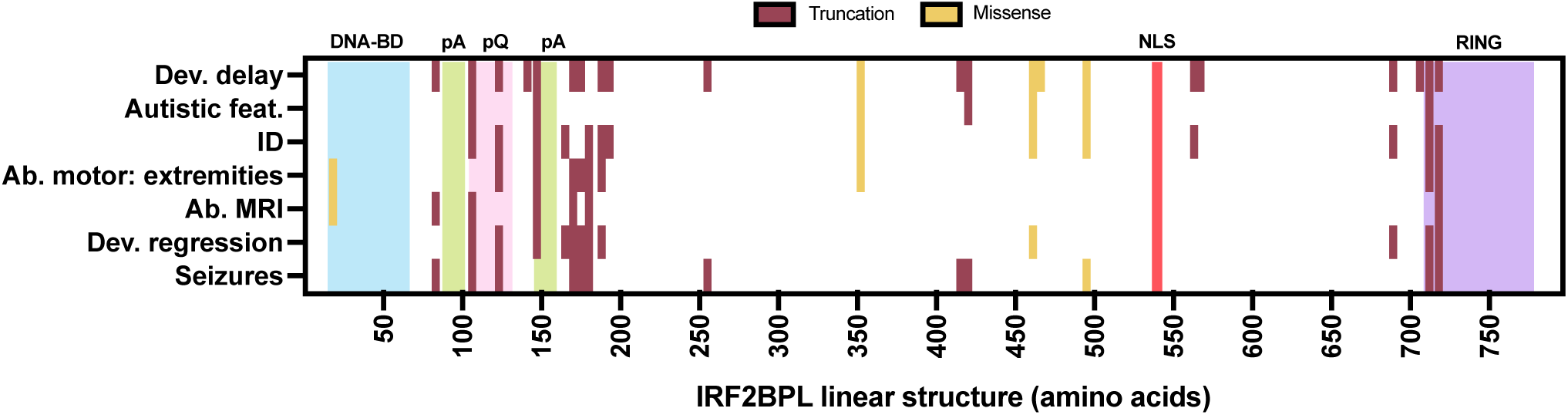
Disease-associated IRF2BPL variants segregate based on truncating vs missense but show wide heterogeneity. IRF2BPL protein structure of 796 amino acids with conserved DNA-binding domain (BD) (light blue), poly Alanine (pA) (green), poly Glutamine (pQ) (pink), nuclear localization signal (NLS) (red), and Really Interesting New Gene (RING) domain (purple). Survey respondents who provided genetic information were mapped onto the protein structure. ID = intellectual disabilities, Ab = abnormal, Dev = developmental. Maroon = truncation (nonsense or frameshift), yellow = missense.

## Discussion

In this retrospective study, parents or guardians of patients with IRF2BPL-related disorder answered an initial survey regarding diagnostic history and current symptoms as well as a series of follow-up surveys regarding neurologic information every six months until 24 months. In this study, patients showed a wide range of non-specific symptoms. This adds to the wide phenotypic spectrum of IRF2BPL-related disorders. Challenges remain to diagnose and acquire pertinent patient information to expand the current knowledge of the disorder.

Of the symptoms reported in patients, DD was the most prominent feature followed by ID and DR. This information is significant as it allows us to characterize key features of IRF2BPL-related disorder. Interestingly, DR was observed in most patients with truncating variants as DR was present in only one patient harboring a missense variant. This supports the hypothesis that *IRF2BPL* missense variants may be less severe than truncating variants.

New seizure and ocular findings were not as common in the six-month follow-up surveys, however new motor abnormalities and speech challenges were more commonly seen. This is important for healthcare providers to be aware of as the onset of symptoms may occur after the initial diagnosis.

Less than half of patients participated long enough to complete the six-month survey and approximately 20% of patients completed the 24-month survey, therefore it is hard to determine if the above conclusions are generalizable to the whole patient population.

Patient-reported data is being used increasingly in research, clinical practice and clinical trial settings. This type of data collection can lead to the acquisition of relevant health information in a timely and cost-effective manner. In addition, a well-developed tool can lead to an increased response rate which leads to a larger study population and potentially increased participant diversity^30^. In the context of an ultra-rare disease such as IRF2BPL-related disorder, other types of data collection may pose a challenge to researchers. Therefore, patient-reported data may be essential in procuring and characterizing phenotypic information for certain diseases.

Limitations in this study include a small patient population which can affect the generalizability of the study as it is difficult to draw conclusions on only 32 participants. In addition to the small patient population, patients with truncating variants were overrepresented as there were 22 patients with truncating variants in comparison to five patients with missense variants. As truncating and missense variants are hypothesized to act differently, it would be important to enroll more patients with missense variants. Furthermore, most of the participants were of European background which is an inaccurate representation of the patient population. Therefore, the study results may be biased towards this population. Although there are benefits to patient-reported data, this type of data collection also provides some limitations such as families falsely or failing to report relevant information. Finally, participant retention throughout the 24 months was a challenge, therefore it is difficult to draw conclusions on follow-up data. However, this initial report is a benchmark for understanding IRF2BPL-related conditions and can serve in designing larger natural history studies and clinical trials. Although there are limitations to patient-reported data collection, this type of research is an accessible tool to help further characterize ultra-rare diseases which can help with overall disease recognition, diagnosis, treatment strategies and increasing the quality of life of patients and their families.

Among the 32 participants in this retrospective study, 27 carried *IRF2BPL* variants, comprising 22 with truncating variants and five with missense variants. While the clinical presentation remains increasingly diverse, common features include developmental delay, intellectual disability, and developmental regression. The findings also support the hypothesis that *IRF2BPL* missense variants are associated with milder symptoms compared to truncating variants. This study contributes to a more comprehensive understanding of IRF2BPL-related disorders, offering valuable clinical and molecular insights into this ultra-rare condition.

## Data Availability

All data produced in the present work are contained in the manuscript

## Acknowledgements

We would like to thank all patients and families that participated in the survey and helped in recruitment across patient-family groups including the Undiagnosed Diseases Network. IRF2BPL studies in PCM lab funded by MMSF (Manitoba Medical Service Foundation), Dystonia Medical Research Foundation Canada, and Rare Diseases: Models & Mechanisms Network (Genome Canada and the Canadian Institutes of Health Research [CIHR]), Research Manitoba and Brian Canada. ZGJ is funded by the CIHR Canada Graduate Scholarships – Master’s program. DMP is funded by the Rady Faculty of Health Sciences PhD Graduate Studentship.

## Author contributions: CRediT

Conceptualization: LDMP; Data curation: LDMP; Formal analysis: LDMP, PCM; Funding acquisition: LDMP, PCM; Methodology: LDMP, PCM; Project administration: LDMP, LAB; Writing – original draft: ZGJ, DMP, PCM; Writing – review and editing: PCM, ZGJ, DMP, LAB, LDMP

## Ethics Statement

This study was reviewed and approved by the Cincinnati Children’s Hospital Institutional Review Board (protocol 2023-0410).

---

**Supplemental Table 1.** List of study participant and associated variants when information was provided.

